# Productivity-adjusted life years lost due to type 2 diabetes by socioeconomic position in Germany between 2022 and 2060

**DOI:** 10.64898/2026.11.20.26368491

**Authors:** Katharina Piedboeuf-Potyka, Ramona Hering, Mandy Schulz, Ralph Brinks, Annika Hoyer, Marielle Wirth, Thaddäus Tönnies

## Abstract

**Introduction:** Type 2 diabetes can impair productivity during working age (20–69 years), causing substantial economic burden at individual and at population level. We aimed to estimate age- and sex-specific productivity-adjusted life years (PALYs) lost due to type 2 diabetes by socioeconomic position (SEP) between 2022 and 2060 in Germany.

**Methods:** We used an established projection model and nationally representative, publicly available German data to estimate age- and sex-specific PALYs lost due to type 2 diabetes by SEP among working-age adults, accounting for future incidence trends. PALYs lost were calculated as the sum of years of life lost (YLL) and years of productivity lost (YPL) due to reduced labour force participation, presenteeism, and absenteeism, using SEP-specific estimates from representative studies.

**Results:** Productivity losses were highest among persons with low SEP, followed by those with medium and high SEP. In 2022, 20-year-old men with type 2 diabetes and low, medium, and high SEP were projected to lose 11.32 PALYs (95% confidence interval (CI) 7.99–14.56), 8.19 PALYs (95% CI 6.01–10.33), and 3.78 PALYs (95% CI 0.89–6.72), respectively, by age 69 compared with men without type 2 diabetes, mainly driven by YPL. A comparable socioeconomic gradient was observed among women and remained largely unchanged until 2060. At the population level, an estimated 9.5 million PALYs (95% CI 6.42–12.58) were lost due to type 2 diabetes in 2022 in both sexes. This number is projected to increase to 9.5–13.5 million by 2060, mainly driven by the projected increase in the number of persons with type 2 diabetes.

**Conclusion:** Type 2 diabetes-related productivity losses were shaped by SEP, sex and the future number of persons with the disease. PALYs lost declined with age and were predominantly driven by YPL.

## Introduction

Type 2 diabetes is associated with several comorbidities, including cardiovascular, neurological, and renal disorders. [1] Beyond physical complications, type 2 diabetes can also impair cognitive function [2] and productivity during working age [3, 4], causing substantial economic burden at both the individual and population level. [5, 6]. Reductions in productivity associated with type 2 diabetes are due to increased mortality, lower productivity while at work (presenteeism), a higher number of sick leave days (absenteeism), and reduced labour force participation (LFP). [7] To quantify the impact of these productivity losses, productivity-adjusted life years (PALYs) have been proposed as a metric that combines mortality and morbidity components into years of productive life lost during working age. [3, 8] Applying this metric, Hird et al. estimated that diabetes resulted in a loss of 75.8 million PALYs over the remaining working lifetime among all persons with diabetes in China in 2017. [9] Similarily, Tönnies et al. projected a population-wide loss of 15.39 million PALYs in 2040 due to type 2 diabetes in Germany. [3]

In light of the substantial projected increase in the number of persons with type 2 diabetes [10] and ongoing demographic changes in Germany which are associated with an aging population and changes in the labour force [3, 6, 11], estimating future productivity losses is essential to inform healthcare planning, policy decisions and persons with type 2 diabetes. Importantly, the burden of type 2 diabetes is not distributed equally across the population. As type 2 diabetes disproportionately affects persons with lower socioeconomic position (SEP) or persons living in deprived regions [12, 13], substantial differences in productivity losses by SEP are likely. Wirth et al. note that persons living in highly deprived regions have a higher prevalence than those living in less deprived regions. [13] Moreover, previous studies indicate that the impact of diabetes on productivity might be stronger among persons in lower SEP. [4, 14]

Hence, this study aims to estimate age- and sex-specific PALYs lost due to type 2 diabetes by SEP between 2022 and 2060.

## Methods

### Calculation of productivity losses by SEP at the individual and population level

We estimated age- and sex-specific productivity losses attributable to type 2 diabetes in Germany between 2022 and 2060 in terms of PALYs lost, assessed at both the individual and population levels. PALYs losses due to type 2 diabetes comprised two components: (1) years of life lost (YLL) due to excess mortality associated with type 2 diabetes and (2) years of productivity lost (YPL) due to diabetes-related morbidity. PALYs lost were calculated as the sum of YLL and YPL. [3]

YLL were defined as the difference in remaining life expectancy between persons with and without type 2 diabetes within the same SEP group, up to the age of 69 years. This upper age limit was chosen to account for the expected increase in the statutory retirement age in Germany from 67 to 69 years over the coming decades. This component captures productivity losses attributable to premature mortality associated with type 2 diabetes. [3]

YPL represented productivity losses due to morbidity among persons with type 2 diabetes and comprised three components: reduced LFP, absenteeism, and presenteeism. These components reflect productivity losses associated with reduced participation in paid employment, absence from work, and reduced productivity while at work, respectively. [3]

Reduced LFP associated with type 2 diabetes was assumed to vary by SEP, based on previous evidence. [4] In addition, persons with type 2 diabetes who participated in the labour force were assumed to have additional absence days per year (absenteeism), with the number of additional days varying by SEP. [4] The SEP-specific values used for both components are provided in the Supplemental Material (Supplement 1). Further details on the calculation of YLL and YPL are provided in Tönnies et al. [3]

**Supplement Table1.** Assumptions for LFP shortfall associated with type 2 diabetes, additional absence days per year [4] and productivity loss due to absenteeism and presenteism [5].

|  | Socioeconomic position |  |  |
|---|---|---|---|
| Productivity loss component | Low | Medium | High |
| LFP shortfall (%) | 17.7 | 11.6 | 3.3 |
| Additional work absence (days/year) | 7.4 | 6.1 | 4.4 |
| Productivity loss due to absenteeism (%) | 3.3 | 2.7 | 1.9 |
| Productivity loss due to presenteeism (%) | 0.5 | 0.5 | 0.5 |

LFP, labour force participation

At the individual level, productivity losses were estimated by comparing a person with type 2 diabetes with a person of the same age (20–69 years), sex (women, men), and SEP (high, medium, low) but without type 2 diabetes over the remaining working life until the age of 69 years. [3] Population-level productivity losses were estimated by summing the individual productivity losses of all persons with type 2 diabetes aged 20–69 years within each year and SEP. [3] This required projections of the age-, sex-, and SEP-specific prevalence and number of persons with type 2 diabetes between 2022 and 2060. [3] The projection model was run until 2109 to enable the estimation of YLL, YPL, and PALYs lost up to the age of 69 years for persons aged 20 years in 2060.

### Projection of prevalence and number of persons with type 2 diabetes by SEP

Future age- and sex-specific type 2 diabetes prevalence and number of persons with type 2 diabetes by SEP was projected using an illness-death model that describes the relationship between incidence (*i*), prevalence (*p*), all-cause mortality (*m*) and the mortality rate ratio (*MRR*) for persons with vs. without type 2 diabetes as functions of age, calendar time and SEP. The temporal change in prevalence (*∂p*) is given by the following partial differential equation (PDE)

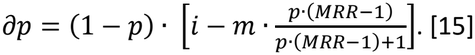

The PDE was solved numerically using nationally representative estimates of prevalence and incidence of type 2 diabetes all-cause mortality, and MRR to project future type 2 diabetes prevalence. Age-, sex-, and SEP-specific prevalence estimates for 2022 from Wirth et al. were used as the baseline prevalence for the projections. [13] Age- and sex-specific type 2 diabetes incidence by SEP for 2022 was estimated by applying age-sex- and SEP-specific, standardized incidence ratios (SIRs) from Piedboeuf-Potyka et al. [12] to national age- and sex-specific incidence estimates from the Central Research Institute for Ambulatory Health Care in Germany (Zi) [16]. Estimates for prevalence and incidence are based on data including all persons in statutory health insurance, which refer to approximately 85% of the German population. [17, 18] SEP-specific all-cause mortality was based on population projections and regional mortality data linked to GISD years. [19, 20] Age- and sex-specific MRR estimates were taken from Schmidt et al. [21]

The projected number of persons with type 2 diabetes was estimated by multiplying the projected age-sex-and SEP-specific prevalence by the corresponding age- and sex-specific population projections from the German Federal Statistical Office. [19] The population projections assumed moderate future changes in life expectancy at birth, fertility, and migration. We only considered the population in working-age from 20–69 years.

### Derivation of the SEP group

The SEP was assessed using the German Index of Socioeconomic Deprivation (GISD). [22] The GISD is a publicly available area-level index based on aggregated regional data and reflects SEP at the county level. [22] It combines information on education, occupation, and income into a score ranging from 0 (highest SEP) to 1 (lowest SEP). [22] Data on incidence, prevalence, and mortality were available by GISD quintile. SEP for absenteeism and reduced LFP was measured at the individual level and categorized into three groups: high, medium, and low SEP. [4] To harmonize the different data sources, the five GISD quintiles were aggregated into the same three SEP categories. The first quintile of the GISD was defined as the high SEP group, quintiles two to four as the medium SEP group, and the fifth quintile as the low SEP group. Ideally, all data sources would use the same SEP indicator. However, we assumed that the different measures were sufficiently comparable, as the GISD was developed based on the indicator used by Mackowiak et al. and serves as a proxy for this measure. [4]

### Probalistic sensitivity analyses and projection Scenarios

We applied three scenarios for the population productivity loss depending on the type 2 diabetes prevalence between 2022 and 2060. Across all projection scenarios, we assumed a constant SEP distribution, represented by the GISD quintile distribution observed in 2019. We further assumed annual reductions in MRRs of 0.60% for women and 0.72% for men with type 2 diabetes, based on evidence from Denmark. [23] As SEP-specific MRR estimates were not available, the age- and sex-specific MRRs reported by Schmidt et al. were applied to all SEP groups. [21] The scenarios differed regarding future trends in age-sex- and SEP-specific type 2 diabetes incidence. [12] In the first Scenario we assume constant type 2 diabetes incidence between 2022 and 2060. In the other scenarios we assume a 5-year de- or increase in type 2 diabetes incidence between 2022 and 2027. Between 2027 and 2060 the incidence remains constant. More details and a visualization of the scenarios can be found in Piedboeuf-Potyka et al. [12]

To account for uncertainty in the input data, the analyses were repeated 1,000 times separately for each scenario, SEP group and sex. In each iteration, input values were randomly sampled from normal distributions parameterized by the corresponding standard errors. The median of the resulting 1,000 estimates for YLL, YPL, and PALYs losses was reported as the point estimate, with the 2.5^th^ and 97.5^th^ percentiles representing the 95% confidence interval (CI).

## Results

### Productivity-adjusted life years lost at the individual level

As future prevalence does not influence results on the individual level, the results in this section are applicable equally to all projection scenarios. Figure 1 shows the expected age-specific loss of productive life years due to type 2 diabetes by sex and SEP in Germany between 2022 and 2060. Across all SEP groups, the expected loss of productive life years decreased with increasing age. A comparable age gradient was observed in both men and women. The expected productivity losses were highest among persons with low SEP, followed by medium and high SEP.

**Figure 1.**
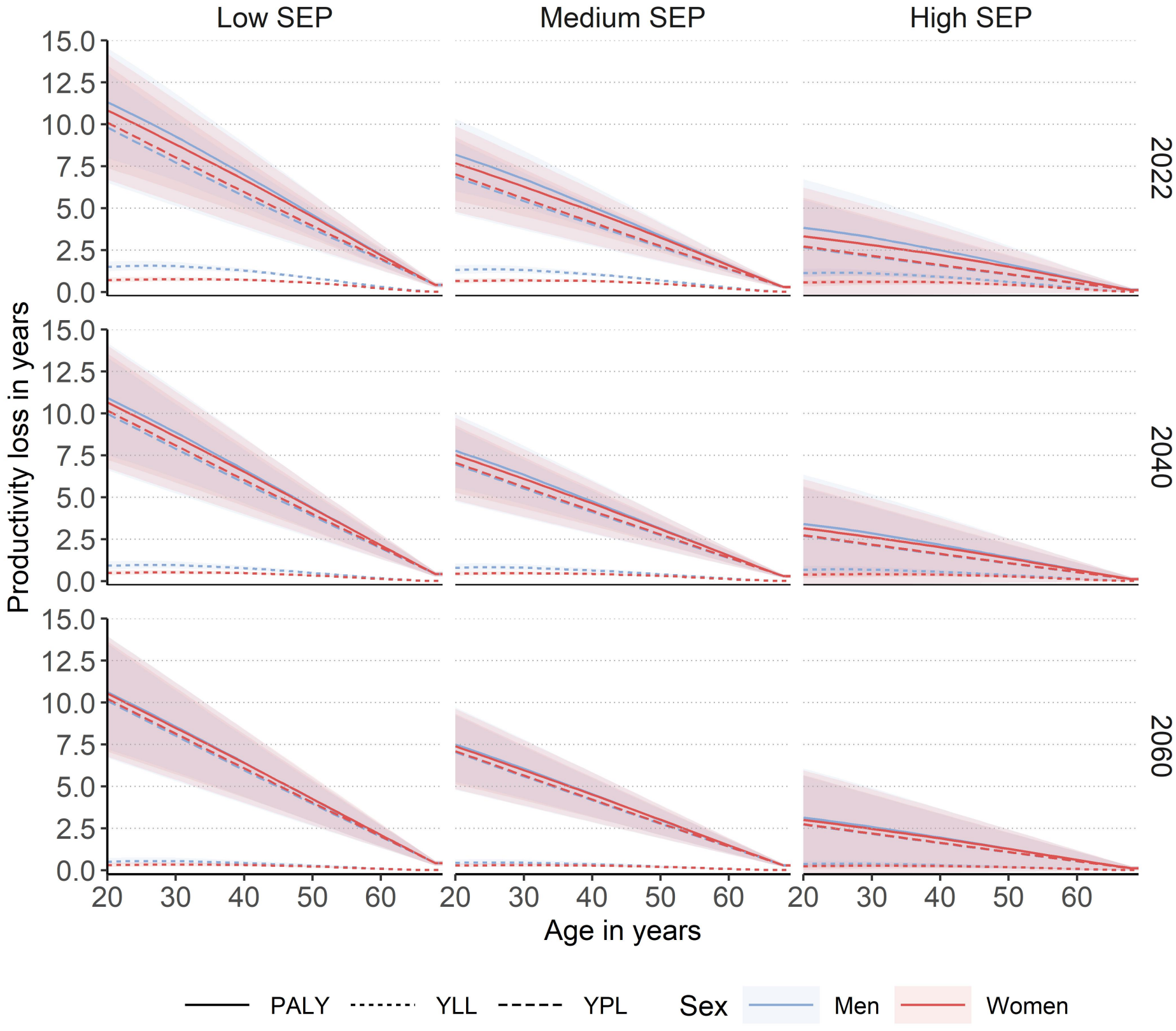
Age-specific productivity adjusted life years (PALYs) lost, years of life lost (YLL) and years of productivity lost (YPL) due to type 2 diabetes by socioeconomic position (SEP) in Germany among men (blue) and women (red) in 2022, 2040 and 2060, Germany. Shaded areas represent 95%-confidence intervals.

In 2022, 20-year-old men with type 2 diabetes and low SEP are expected to lose 11.3 PALYs (95% CI 8.3–14.7) up to age 69 compared to men without type 2 diabetes, whereas those with high SEP are expected to lose 3.8 PALYs (95% CI 1.1–6.7) (Table 1). At age 69, men with low SEP are expected to lose 0.42 PALYs (95% CI 0.28–0.56), compared with 0.12 PALYs (95% CI 0.00–0.23) among those with high SEP (Table 1). A comparable socioeconomic gradient was observed among women. Across all SEP groups, productivity losses attributable to type 2 diabetes declined slightly between 2022 and 2060 (Table 1).

**Table 1.** Expected loss of productive life years of 20- and 69-year-oldmen and women in 2022, 2040 and 2060, Germany.

|  |  |  | Age = 20 years |  |  | Age = 69 years |  |  |
|---|---|---|---|---|---|---|---|---|
| Year | Sex | SEP | PALYs (95%-CI) | YPL (95%-CI) | YLL (95%-CI) | PALYs (95%-CI) | YPL (95%-CI) | YLL (95%-CI) |
| 2022 | Women | Low | 10.84 (7.40–14.20) | 10.11 (6.70–13.51) | 0.71 (0.57–0.87) | 0.42 (0.28–0.56) | 0.41 (0.27–0.55) | 0.01 (0.01–0.01) |
|  |  | Medium | 7.69 (5.47–9.90) | 7.03 (4.79–9.24) | 0.64 (0.52–0.80) | 0.29 (0.20–0.38) | 0.28 (0.19–0.37) | 0.01 (0.01–0.01) |
|  |  | High | 3.32 (0.33–6.23) | 2.72 (-0.24–5.63) | 0.56 (0.45–0.70) | 0.12 (0.00–0.24) | 0.11 (-0.01–0.23) | 0.01 (0.01–0.01) |
|  | Men | Low | 11.32 (7.99–14.56) | 9.82 (6.52–13.12) | 1.50 (1.23–1.82) | 0.42 (0.28–0.56) | 0.41 (0.27–0.54) | 0.01 (0.01–0.01) |
|  |  | Medium | 8.19 (6.01–10.33) | 6.87 (4.68–9.01) | 1.31 (1.08–1.60) | 0.29 (0.20–0.38) | 0.28 (0.19–0.37) | 0.01 (0.01–0.01) |
|  |  | High | 3.78 (0.89–6.72) | 2.66 (-0.24–5.52) | 1.12 (0.92–1.38) | 0.12 (0.00–0.24) | 0.11 (-0.01–0.11) | 0.01 (0.01–0.01) |
| 2040 | Women | Low | 10.66 (7.24–14.06) | 10.17 (6.74–13.59) | 0.48 (0.36–0.63) | 0.42 (0.28–0.55) | 0.41 (0.27–0.55) | 0.01 (0.01–0.01) |
|  |  | Medium | 7.53 (5.28–9.75) | 7.07 (4.82–9.29) | 0.44 (0.32–0.58) | 0.29 (0.20–0.38) | 0.28 (0.19–0.37) | 0.01 (0.00–0.01) |
|  |  | High | 3.15 (0.17–6.08) | 2.73 (-0.25–5.66) | 0.38 (0.28–0.51) | 0.12 (0.00–0.23) | 0.11 (-0.01–0.23) | 0.01 (0.00–0.01) |
|  | Men | Low | 10.93 (7.52–10.93) | 9.98 (6.62–13.35) | 0.92 (0.70–1.19) | 0.42 (0.28–0.55) | 0.41 (0.27–0.55) | 0.01 (0.01–0.01) |
|  |  | Medium | 7.78 (5.58–9.97) | 6.97 (4.75–9.15) | 0.80 (0.60–1.04) | 0.29 (0.20–0.38) | 0.28 (0.19–0.37) | 0.01 (0.01–0.01) |
|  |  | High | 3.41 (0.44–6.35) | 2.70 (-0.24–5.59) | 0.68 (0.51–0.89) | 0.12 (0.00–0.23) | 0.11 (-0.01–0.23) | 0.01 (0.01–0.01) |
| 2060 | Women | Low | 10.54 (7.08–13.95) | 10.22 (6.77–13.66) | 0.31 (0.20–0.45) | 0.42 (0.28–0.55) | 0.41 (0.27–0.55) | 0.00 (0.00–0.00) |
|  |  | Medium | 7.39 (5.14–9.63) | 7.10 (4.84–9.33) | 0.28 (0.18–0.41) | 0.29 (0.20–0.38) | 0.29 (0.19–0.38) | 0.00 (0.00–0.00) |
|  |  | High | 3.00 (0.02–5.96) | 2.75 (-0.25–5.68) | 0.24 (0.15–0.36) | 0.11 (-0.01–0.23) | 0.11 (-0.01–0.23) | 0.00 (0.00–0.00) |
|  | Men | Low | 10.63 (7.22–13.98) | 10.10 (6.70–13.51) | 0.51 (0.35–0.74) | 0.41 (0.28–0.55) | 0.41 (0.27–0.55) | 0.00 (0.00–0.01) |
|  |  | Medium | 7.50 (5.27–9.71) | 7.04 (4.80–9.25) | 0.44 (0.30–0.64) | 0.29 (0.20–0.38) | 0.29 (0.19–0.38) | 0.00 (0.00–0.00) |
|  |  | High | 3.14 (0.16–6.07) | 2.73 (-0.25–5.64) | 0.37 (0.26–0.54) | 0.11 (-0.01–0.23) | 0.11 (-0.01–0.23) | 0.00 (0.00–0.00) |

SEP, socioeconomic position; PALYs, productivity adjusted life years; YPL, years of productivity lost; YLL, years of life lost; CI, confidence interval.

Figure 1 and Table 1 show that productivity losses caused by presenteeism, absenteeism and reduced labour force (YPL) was the predominant contributor to PALYs losses, exceeding the contribution of YLL across all SEP groups, sexes, and projection years.

### Productivity life years lost at the population level

Figure 2 shows the expected PALYs loss at population level due to type 2 diabetes in Germany between 2022 and 2060 for all three scenarios by sex.

**Figure 2.**
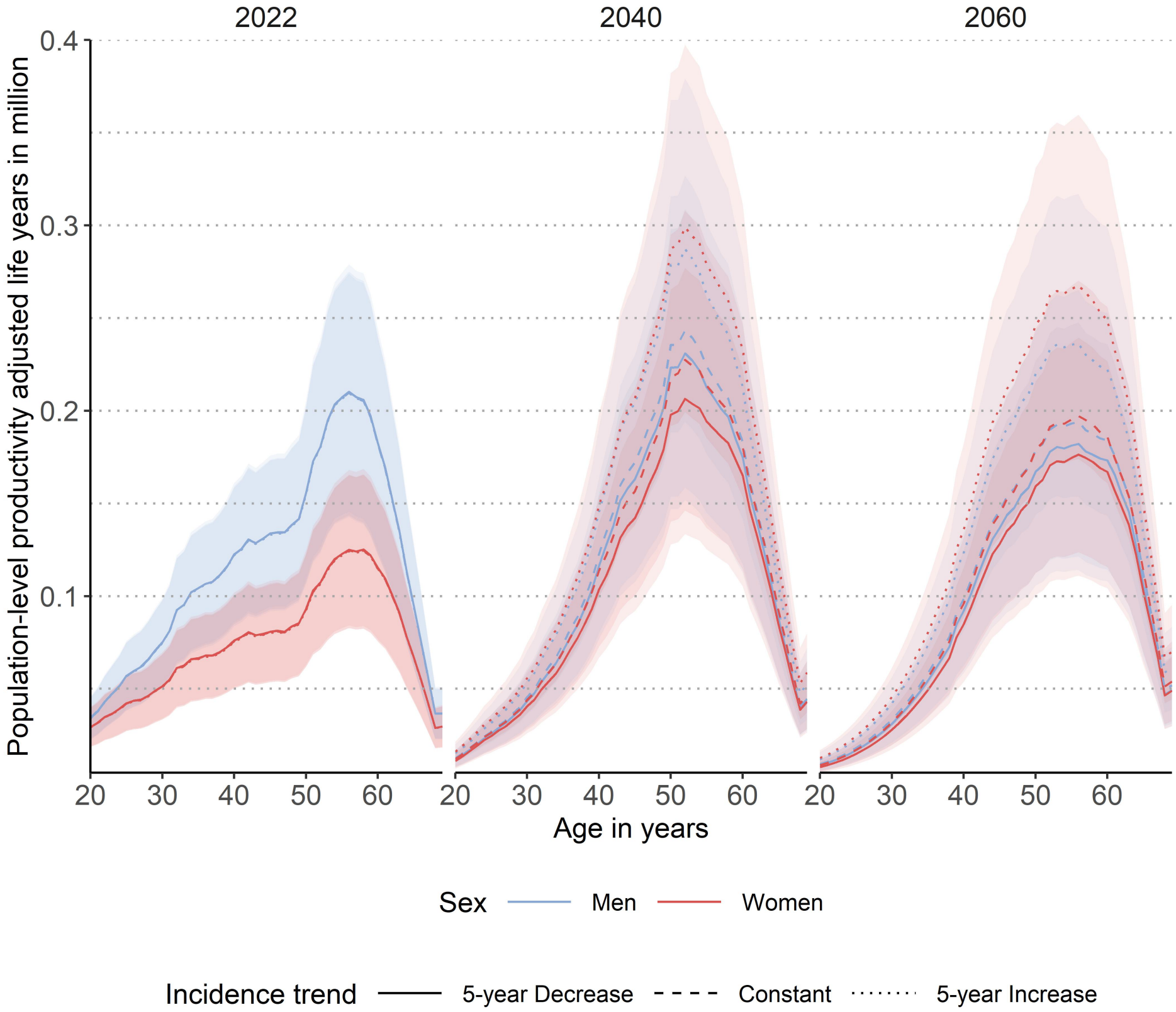
Population-level age-specific productivity adjusted life years (PALYs) lost due to type 2 diabetes in Germany among men (blue) and women (red) between 2022 and 2060, shown for all three applied incidence trends. Decreasing and increasing scenarios show declining or rising type 2 diabetes incidence until 2027, respectively, while the constant scenario assumes stable incidence from 2022 to 2060. Scenarios with increasing type 2 diabetes incidence extrapolate the non-linear trends observed between 2014 and 2022, capturing the recent increase, whereas decreasing scenarios extrapolate the long-term linear decrease in incidence. Shaded areas represent 95%-confidence intervals.

In each projection year, population-level PALYs losses are closely linked to type 2 diabetes prevalence, increasing with age and peaking between the ages of 50 and 60.

Between 2022 and 2040, PALYs losses increase, followed by a decline by 2060. Differences between the incidence scenarios are more pronounced than those between projection years. PALYs losses are lowest under the scenario assuming a 5-year decline in incidence and highest under the scenario assuming a 5-year increase in incidence. Sex differences in PALYs losses are most pronounced in 2022 but become less apparent in 2040 and 2060.

In 2022, approximately 4 (95% CI 3.88-4.19) million persons in working age (20–69 years) were living with type 2 diabetes (Table 2). By 2040, these numbers are expected to increase to between 4.93 (95% CI 4.83–5.02) and 6.43 (95% CI 6.31–6.56) million, and to between 5.04 (95% CI 4.93–5.14) and 6.94 (95% CI 6.79–7.08) million by 2060, depending on the trend of future incidence.

**Table 2.** Number of persons with type 2 diabetes in Germany between 2022 and 2060 by sex.

|  |  | Number of persons with type 2 diabetes in million |  |  |
|---|---|---|---|---|
| Year | Sex | 5-year decrease<br>(95%-CI) | Constant incidence<br>(95%-CI) | 5-year increase<br>(95%-CI) |
| 2022 | Women | 1.59 (1.52–1.66) | 1.59 (1.52–1.66) | 1.59 (1.52–1.66) |
|  | Men | 2.45 (2.37–2.53) | 2.45 (2.37–2.53) | 2.45 (2.37–2.54) |
|  | **Total** | **4.04 (3.89–4.19)** | **4.04 (3.88–4.19)** | **4.04 (3.89–4.19)** |
| 2040 | Women | 2.34 (2.30–2.39) | 2.56 (2.51–2.61) | 3.30 (3.24–3.37) |
|  | Men | 2.58 (2.54–2.63) | 2.70 (2.65–2.75) | 3.13 (3.07–3.19) |
|  | **Total** | **4.93 (4.83–5.02)** | **5.26 (5.16–5.36)** | **6.43 (6.31–6.56)** |
| 2060 | Women | 2.45 (2.40–2.50) | 2.73 (2.67–2.78) | 3.65 (3.58–3.73) |
|  | Men | 2.59 (2.53–2.64) | 2.73 (2.68–2.79) | 3.28 (3.21–3.35) |
|  | **Total** | **5.04 (4.93–5.14)** | **5.46 (5.35–5.58)** | **6.94 (6.79–7.08)** |

CI, confidence interval.

These estimates corresponded to substantial productivity losses (Table 3). In 2022, estimated PALYs lost were similar across all three scenarios, ranging from 9.51 (95% CI 6.51– 12.54) to 9.55 (95% CI 6.42– 12.58) million PALYs. By 2040, the PALYs lost were estimated to increase to 10.84 million (95% CI 7.02– 14.55) under the 5-year decrease scenario, 11.63 million (95% CI 7.47– 15.72) in the constant-incidence scenario, and 14.38 million (95% CI 9.60– 19.11) in the 5-year increase scenario. By 2060, PALYs lost were projected to reach 9.54 million (95% CI 5.99– 12.96), 10.42 million (95% CI 6.52– 14.29), and 13.55 million (95% CI 8.81– 18.19), respectively.

**Table 3.** Population level productivity adjusted life years (PALYs) lost due to years of life lost (YLL), and years of productivity lost (YPL) among persons with type 2 diabetes of working age between 2022 and 2060 in Germany.

| Year | Sex | 5-year decrease (95%-CI) |  |  | Constant incidence (95%-CI) |  |  | 5-year increase (95%-CI) |  |  |
|---|---|---|---|---|---|---|---|---|---|---|
|  |  | PALYs | YPL | YLL | PALYs | YPL | YLL | PALYs | YPL | YLL |
| 2022 | Women | 3.72 (2.44–4.94) | 3.23 (1.99–4.45) | 0.48 (0.43–0.54) | 3.71 (2.43–5.01) | 3.22 (1.97–4.54) | 0.48 (0.42–0.54) | 3.69 (2.48–4.92) | 3.23 (2.01–4.45) | 0.47 (0.42–0.52) |
|  | Men | 5.83 (3.97–7.64) | 4.72 (2.89–6.51) | 1.12 (1.00–1.23) | 5.82 (3.94–7.74) | 4.71 (2.86–6.63) | 1.11 (0.99–1.23) | 5.81 (4.03–7.61) | 4.71 (2.94–6.49) | 1.09 (0.99–1.21) |
|  | **Total** | **9.55 (6.42–12.58)** | **7.95 (4.88–10.96)** | **1.59 (1.43–1.77)** | **9.53 (6.38–12.76)** | **7.94 (4.83–11.17)** | **1.59 (1.41–1.76)** | **9.51 (6.51–12.54)** | **7.94 (4.95–10.94)** | **1.56 (1.42–1.74)** |
| 2040 | Women | 5.16 (3.32–6.96) | 4.69 (2.84–6.46) | 0.48 (0.39–0.56) | 5.66 (3.61–7.69) | 5.14 (3.10–7.20) | 0.52 (0.43–0.61) | 7.37 (4.89–9.83) | 6.74 (4.24–9.18) | 0.64 (0.54–0.74) |
|  | Men | 5.67 (3.70–7.59) | 4.99 (3.02–6.88) | 0.69 (0.56–0.82) | 5.96 (3.86–8.03) | 5.23 (3.15–7.34) | 0.73 (0.59–0.87) | 7.01 (4.71–9.27) | 6.17 (3.87–8.41) | 0.83 (0.69–0.99) |
|  | **Total** | **10.84 (7.02–14.55)** | **9.68 (5.87–13.35)** | **1.17 (0.95–1.39)** | **11.63 (7.47–15.72)** | **10.38 (6.26–14.54)** | **1.24 (1.02–1.48)** | **14.38 (9.6–19.11)** | **12.92 (8.12–17.60)** | **1.47 (1.23–1.74)** |
| 2060 | Women | 4.64 (2.92–6.31) | 4.37 (2.64–6.04) | 0.27 (0.19–0.36) | 5.21 (3.25–7.15) | 4.90 (2.94–6.86) | 0.30 (0.21–0.41) | 7.14 (4.64–9.60) | 6.75 (4.25–9.19) | 0.39 (0.29–0.52) |
|  | Men | 4.89 (3.08–6.65) | 4.58 (2.77–6.32) | 0.31 (0.20–0.45) | 5.21 (3.26–7.14) | 4.87 (2.92–6.83) | 0.34 (0.22–0.49) | 6.40 (4.17–8.59) | 5.98 (3.77–8.15) | 0.41 (0.27–0.58) |
|  | **Total** | **9.54 (5.99–12.96)** | **8.95 (5.41–12.36)** | **0.59 (0.39–0.81)** | **10.42 (6.52–14.29)** | **9.77 (5.87–13.69)** | **0.65 (0.42–0.89)** | **13.55 (8.81–18.19)** | **12.74 (8.02–17.35)** | **0.81 (0.56–1.09)** |

Decreasing and increasing scenarios show declining or rising type 2 diabetes incidence until 2027, respectively, while the constant scenario assumes stable incidence from 2022 to 2060. Scenarios with increasing type 2 diabetes incidence extrapolate the non-linear trends observed between 2014 and 2022, capturing the recent increase, whereas decreasing scenarios extrapolate the long-term linear decrease in incidence.
CI, confidence interval.

## Discussion

Using an illness-death model, we projected future age- and sex-specific PALYs lost due to type 2 diabetes by SEP, at both the individual and population level in Germany between 2022 and 2060. The key novel finding of this study is the substantial socioeconomic gradient in future productivity losses associated with type 2 diabetes. Our results show that persons with lower SEP are expected to experience considerably greater PALYs lost than those with higher SEP, reflecting differences in both years of potential life lost and productivity losses. By incorporating SEP into projections of the future burden of type 2 diabetes, our study highlights that the economic consequences of type 2 diabetes are likely to be distributed unequally across socioeconomic groups.

At the individual level, a 20-year-old men and a woman of the same age in the low SEP group are projected to lose approximately three times as many PALYs as their counterparts in the high SEP group. At the population level, future trends in type 2 diabetes incidence are a main determinant of future productivity losses. A moderate increase in incidence over five years resulted in the highest PALYs lost, followed by a constant incidence and a five-year decline in incidence. The differences in PALYs lost were smaller among persons with medium SEP and virtually nonexistent among those with high SEP.

### Comparison with previous studies

To our knowledge, no previous studies projected productivity losses associated with type 2 diabetes while accounting for SEP. Although our overall estimates can be compared with previous studies, the lack of SEP-specific analyses in the existing literature precludes direct comparisons of the socioeconomic gradient in productivity losses. Our study thus adds a novel perspective by demonstrating how the future productivity burden of type 2 diabetes may differ across socioeconomic groups. Tönnies et al. also reported the highest individual productive life year losses among younger adults with type 2 diabetes in Germany, with women aged 20 years expected to lose 12.8 PALYs compared with women without type 2 diabetes. [3] These losses declined with increasing age. At the population level, they projected an increase in total PALYs lost from 12.06 million in 2020 to 15.39 million in 2040, primarily driven by productivity-related losses (YPL), which accounted for 10.44 million and 14.06 million PALYs lost, respectively. This is consistent with our finding that YPL contributed more to total PALYs lost than YLL. This increase in total PALYs lost was accompanied by a projected rise in the number of persons with type 2 diabetes from 4.60 million in 2020 to 5.45 million in 2040, as estimated by Tönnies et al. [3] Magliano et al. estimated substantial PALYs lost attributable to diabetes in an Australian population using a different methodological approach to PALYs estimation than the one applied in our study. [8] Despite these methodological differences, their findings were qualitatively consistent with ours, showing greater PALYs lost among younger than older persons and demonstrating that diabetes reduced both years of life lived and productive years of life lived. Evidence from China also demonstrates a substantial impact of diabetes on PALYs, highlighting that diabetes-related productivity losses represent a relevant burden across different populations. [9] Extending these findings, our study demonstrates pronounced socioeconomic inequalities, with persons of low SEP consistently experiencing the greatest productivity losses. These differences may partly reflect socioeconomic differences in occupational structures, as persons in lower SEP groups are more likely to work in physically demanding occupations, such as construction and manual labour, where working despite illness may be more difficult than in office-based occupations.

### Implications for future research and public health

Our findings highlight socioeconomic inequalities in productivity losses associated with type 2 diabetes, with persons with low SEP facing a double burden of a higher prevalence of diabetes and greater productivity losses among those affected by the disease.

Given demographic changes and the economic burden of type 2 diabetes at both the individual and population levels, these findings have important implications for health policy. To reduce the future burden of productivity losses, effective strategies for the prevention and treatment of type 2 diabetes are needed, supplemented by measures to reduce the socioeconomic inequalities that contribute to these disparities.

Since the incidence of type 2 diabetes is a key factor for future prevalence, one approach would be to reduce the future incidence of type 2 diabetes. [10, 12, 24] Measures to reduce or delay the onset of the disease primarily include lifestyle changes, such as increased physical activity or lower-calorie diets aimed at reducing body fat. [25] In addition, high-risk patients (persons with an HbA1c of 6.0%– 6.4% and persons with overweight) can be identified early by the healthcare system and supported through prevention programs or technology-based programs. [25, 26] Gestational diabetes is another important risk factor for type 2 diabetes. Women with a history of gestational diabetes have a substantially higher risk of developing type 2 diabetes, highlighting the importance of effective prevention strategies during pregnancy and the postpartum period. [25]

As persons with low SEP are at increased risk of developing type 2 diabetes, it is essential that preventive interventions effectively reach these populations. [12] Reducing barriers to participation and ensuring that prevention programmes are accessible and tailored to their needs may improve their effectiveness and contribute to reducing health inequalities.

Beyond strategies aimed at preventing type 2 diabetes, measures to reduce diabetes-related complications and improve diabetes management may help mitigate the adverse consequences of the disease, including productivity losses. [27] In particular, early detection and appropriate management of complications may help preserve persons’ health and work ability, thereby reducing diabetes-related productivity losses.

### Strength and limitations

The key strength of our study is the illness-death model we used, which allows to project future productivity losses at both the individual and population level in dependence of SEP. This enables the quantification of socioeconomic inequalities across SEP groups and highlighting the particularly high future burden among socioeconomically disadvantaged populations. Moreover, the PALYs measure integrates various determinants of productivity, such as mortality, presenteeism, absenteeism, and LFP, into a single number. The use of the time scale facilitates the intuitive interpretation of productivity losses and supports the communication of our results to decision makers and persons with type 2 diabetes. Including the SEP in the model provides decision makers with a basis for targeting prevention and care measures at population groups with the greatest need.

However, the projections relied on several assumptions and input parameters that varied in the strength of the available evidence. Moreover, the maximum projection horizon extended to 2109, requiring assumptions about future trends over several decades and thereby introducing additional uncertainty. For age- and sex-specific type 2 diabetes prevalence and incidence, as well as the MRR associated with diabetes, we relied on published estimates based on data from German statutory health insurance covering approximately 85% of the German population [12, 13, 21]. These sources provided precise input values but had the drawback that they took into account only diagnosed cases of diabetes that were documented in clinical practice for reimbursement purposes. Persons who are privately insured or uninsured are also not included in the data. For temporal trends in the MRR, we had to rely on evidence from Denmark due to the lack of evidence from Germany. [23] The use of German-specific MRR trends could have improved the accuracy of the projections. Moreover, the assumptions regarding the annual change in MRR were not differentiated by SEP. However, this limitation is expected to have only a limited impact on our future prevalence projections, given that incidence has a considerably greater influence on future prevalence than mortality. [10, 24] Nevertheless, it may have a greater impact on the projected PALYs, particularly on YLL, as these estimates directly depend on mortality.

Input data for absenteeism and reduced LFP [4] were not available by sex which would likely have improved our projections, since previous studies have reported sex differences in diabetes-related LFP [5]. The LFP shortfall in high income countries (HICs) was assumed to be 12.6% for men and 25.2% for women. Likewise, input data for presenteeism was assumed equal across age, sex and SEP due to the absence of more detailed previuos studies.

In addition, our projections assume that general economic trends remain constant over time. The SEP is based on the 2019 quintile distribution and was assumed to remain constant throughout the observation period. Consequently, potential future changes in the socioeconomic distribution were not considered. Moreover, since the GISD is an area-level measure of SEP, it may not fully reflect individual socioeconomic circumstances and probably underestimates inequalities on the individual level.

To facilitate linkage with other data sources, we aggregated the original five GISD quintiles into three SEP groups (low, medium, and high SEP). For the absenteeism and LFP estimates, SEP was defined using an individual-level index [4] which already distinguished between low, medium, and high SEP. Harmonisation of the GISD quintiles into three SEP groups was therefore necessary to ensure consistent SEP categorisation across data sources and enable their integration into the projections. This broader categorisation may have resulted in some loss of information regarding socioeconomic differences, particularly within the medium-SEP group, which combines three original GISD quintiles. However, as the lowest and highest SEP quintiles were retained unchanged, the contrast between the extremes of the socioeconomic distribution was preserved. The main limitation is therefore a reduced ability to capture socioeconomic gradients and differences between adjacent SEP groups.

## Conclusion

Future productivity losses will be shaped by SEP, sex, and the future trajectory of type 2 diabetes incidence. If current incidence trends persist, socioeconomic inequalities in productivity losses are expected to widen, resulting in a disproportionate burden among socioeconomically disadvantaged populations. To prevent further increases in socially determined health inequalities, prevention and health promotion strategies should prioritize population groups with low SEP.

## Funding

Funded by the Deutsche Forschungsgemeinschaft (DFG, German Research Foundation) – project number 516146656.

## Conflicts of Interest

The authors declare no conflicts of interest.

## Acknowledgements

We thank the Central Research Institute for Ambulatory Health Care in Germany (Zi) for sharing data used in this publication and for their support.

AI was used to shorten and to optimize text passages.

## Data Availability

The GISD data and a documentation of the data are public available under the following link: https://doi.org/10.5281/zenodo.6840304.

## Ethics approval

This study was conducted in accordance with the principles of the Declaration of Helsinki. The Ethics Committee of the Medical Faculty of Heinrich Heine University Düsseldorf has approved this study (study number: 2024-2722).

Only data that is routinely documented for billing purposes in the context of outpatient care, that has already been published, or that is made publicly available by the federal and state statistical offices will be used.

No individual data is available for this research project, meaning that no individuals can be identified. Consequently, no consent can be obtained from individuals.

## Supplemental Material

The labour force participation (LFP) shortfall associated with type 2 diabetes was assumed to be 17.7%, 11.6%, and 3.3% for persons with low, medium, and high SEP, respectively, based on previous evidence. [4] In addition, persons with type 2 diabetes participating in the labour force were assumed to have additional 7.4, 6.1, and 4.4 days of work absence per year (absenteeism) for persons with low, medium, and high SEP, respectively, based on previous evidence. [4]

Assuming 255 working days and 31 paid leave days per year in Germany [28], these estimates correspond to additional annual productivity losses due to absenteeism of 3.3%, 2.7%, and 1.9% for persons with low, medium, and high SEP, respectively. Following Bommer et al., productivity while at work (presenteeism) was assumed to be 0.5% lower among persons with type 2 diabetes than among those without the disease, regardless of SEP. [5]

The percentage of productivity lost due to absenteeism and presenteeism was combined into a productivity loss weight for each SEP group by summing the contributions of both components. All assumptions are summarized in Supplement 1.

## Notes

### Competing Interest Statement

The authors have declared no competing interest.

### Author Declarations

This study was conducted in accordance with the principles of the Declaration of Helsinki. The Ethics Committee of the Medical Faculty of Heinrich Heine University Duesseldorf has approved this study (study number: 2024-2722). Only data that is routinely documented for billing purposes in the context of outpatient care, that has already been published, or that is made publicly available by the federal and state statistical offices will be used. No individual data is available for this research project, meaning that no individuals can be identified. Consequently, no consent can be obtained from individuals.

